# Comparison of Carotid Intima Media Thickness between Women with History of Preeclampsia and Normal Pregnancy: A Meta-Analysis of Systematic Review

**DOI:** 10.1101/2021.01.24.21250392

**Authors:** Budi Susetyo Pikir, Agus Subagjo, Deasy Eka Wardhani, Andrianto, Yudi Her Oktaviono, Ricardo Adrian Nugraha

**Author notes:** Corresponding Author: Prof. Budi Susetyo Pikir, M.D., Ph.D., Department of Cardiology and Vascular Medicine, Faculty of Medicine Universitas Airlangga - Dr. Soetomo General Academic Hospital, Surabaya - INDONESIA, Corresponding Address : Gedung PPJT Lantai 7, Dr. Soetomo General Academic Hospital, Jalan Mayjend Prof. Dr. Moestopo 6-8, Surabaya – INDONESIA, Corresponding.

## Abstract

**Background:** Women with a history of preeclampsia are twice as likely to experience long term cardiovascular disease (CVD) compared to women with unaffected pregnancy. The pathophysiology of preeclampsia is not well understood, however there is general agreement that, similar to cardiovascular disease, endothelial dysfunction plays a crucial role. On a clinical level, preeclampsia and atherosclerotic cardiovascular disease share common risk factors. Carotid intima media thickness (CIMT) is ultrasound-based imaging, non-invasive, simple and reproducible method of subclinical atherosclerosis evaluation. Nowadays, there were studies concerning of CIMT among preeclamptic women, although the results were different.

**Objective:** To prove that CIMT among women with histories of preeclampsia was greater compared to normal pregnancy.

**Methods:** We conducted a meta-analysis of studies that reported CIMT, in women who had preeclampsia and had normal pregnancy. Studies were identified through three databases: PubMed, Google Scholar dan SAGE Journals with publication year of 2010- 2020. Heterogeneity was assessed using the *I*^*2*^ statistic. Standardized mean difference was used as measured of effect size.

**Results:** Nine eligible studies were included in the meta-analysis. This meta-analysis consisted of 439 women with preeclampsia histories and 526 women with normal pregnancy histories. Women who had preeclampsia had significantly higher CIMT compared to those with normal pregnancy with standardized mean difference −0.38 and 95% confidence interval (CI) −0.68 to −0.07 (*p*=0.02).

**Conclusion:** CIMT was greater among women with histories of preeclampsia compared to normal pregnancy.

**Prospero registration number:** ID 228825.

## INTRODUCTION

Preeclampsia is a heterogeneous disease which can be partially described by the presence of uteroplacental and maternal vasculopathy.^1^ Preeclampsia is associated with high maternal and infant morbidity and mortality, which is largely due to dysfunction of the cardiovascular and cerebrovascular systems.^2^

Women with a history of preeclampsia have at least a twofold increased risk of developing various manifestations of cardiovascular disease. The risk of developing chronic hypertension in particular was four times that of women with a history of pregnancy without preeclampsia. In addition, several meta-analyses have shown a 2-3-fold increased risk of developing ischemic heart disease, cerebrovascular disease by 2 times, and heart failure by 4-fold in preeclampsia.^3^ A retrospective cohort study involving 1.6 million pregnancies demonstrated the same, namely that hypertensive disease in pregnancy is associated with premature cardiovascular disease in young women. In tracing 2.7 years after pregnancy, women with preeclampsia had a 2.5 times increased risk of myocardial infarction, 3 times increased risk of heart failure, and 2.3 times increased risk of stroke, compared to women without preeclampsia.^4^

The cellular and molecular mechanisms underlying preeclampsia are not well understood.^5^ However, there is general agreement that, like cardiovascular disease, endothelial dysfunction plays an important role in the pathogenesis of preeclampsia. And at a clinical level, preeclampsia and atherosclerotic cardiovascular disease share the same major risk factors. The existence of advanced complications of preeclampsia such as ischemic heart disease, stroke, and heart failure further supports the idea that preeclampsia is a condition associated with an atherosclerotic process.^3^ This clinical observation is also supported by the presence of histological studies of placental vascular changes in women with preeclampsia. There is acute athetoses of the placental vessels in preeclamptic women, which consists of foam cells containing subendothelial fat, fibrinoid necrotic tissue from the arterial walls, and perivascular lymphocytic infiltration, similar to that found in early stage atherosclerosis.^6^

Angiography and Doppler examinations have been used for more than 30 years to determine the presence of atherosclerosis and its progression, but evaluation is limited to severe atherosclerosis, where stenosis has occurred.^7^ In the late 1980s, advances in ultrasound resolution techniques provided an opportunity to assess and evaluate the atherosclerosis process from precursor lesions to the stage of stenosis using non-invasive techniques.^8^ Carotid intima-media thickness (CIMT) is a method used to assess the presence of subclinical atherosclerosis by combining the intima and media layer thickness components of the artery wall, which is carried out using a non-invasive examination, namely ultrasonography-based imaging.^6^

### OBJECTIVE

To review the comparison of CIMT value between women with a history of preeclampsia and normal pregnancies. Hereby, we described the comparison of classic cardiovascular risk factors between women with a history of preeclampsia and normal pregnancies, and described the comparison of specific biomarkers of atherosclerosis between women with a history of preeclampsia and normal pregnancies.

## METHODS

### Inclusion Criteria

The inclusion criteria were articles derived from studies comparing CIMT between women with a history of preeclampsia and normal pregnancies. The exclusion criteria were research reports other than English language, published before 2010, study data are not in mean-SD form, and CIMT examinations other than in the common carotid artery segment.

### Search Strategy

Article searches were carried out through the electronic database PubMed, Google Scholar and SAGE Journals with the publication year 2010-2020. The key word used in the search is a combination of: (preeclampsia OR pre-eclampsia) AND “without preeclampsia” OR “normal pregnancies” OR “normal pregnant” OR “normotensive pregnancies” OR “normotensive pregnant” OR “uncomplicated pregnancies” AND (“carotid intima media thickness “OR” carotid intima-media thickness “OR CIMT).

### Study Validity Assessment

We evaluated the quality of the studies included, in terms of allocation concealment, blinding of investigators, participants and outcome assessors and completeness of follow-up. Eligibility and study validity assessment was performed independently in an unblinded standardized manner by six reviewers

### Data Extraction

Data extraction was performed on full-text copies of all included studies by the six reviewers independently, using data extraction forms developed for this purpose. Data were extracted from all included studies in terms of preeclampsia and CIMT assessment. Any additional information required from original investigators was requested by written correspondence, and if relevant information was obtained in this manner, this was included in the review. If an outcome was reported at more than one time point for a single study, the longest period of follow-up was used. To avoid data duplication and double data counting, the following were double checked in the databases used: author names, journal names and issues, titles and publications, and date of potentially eligible studies.

### Statistical Analysis

All articles that meet the research requirements are collected and processed using a computer. Statistical analysis was performed using Stata 16 software. This study had a significance level (α) of 0.05 or 5%. Heterogeneity (*I*^*2*^) was used to determine the discrepancy for each CIMT examination. The random effect model is used in this study due to the high level of heterogeneity.

## RESULTS

### Trial Flow and Study Characteristic

The combined search of PubMed, Google Scholar and SAGE Journals, which also included some hand-searching of relevant cardiology and obstetrics journals, retrieved 1,073 articles. After discarding a number of duplicates retrieved by individual searches and reviewing all titles and abstracts, many studies were excluded because they were not relevant with our topics, or did not investigate any of the outcomes of interest to this study, or were animal or basic research studies or review articles. Overall, nine studies, enrolling a total of 965 patients, were included in this analysis (Figure 1).

**Figure 1.**
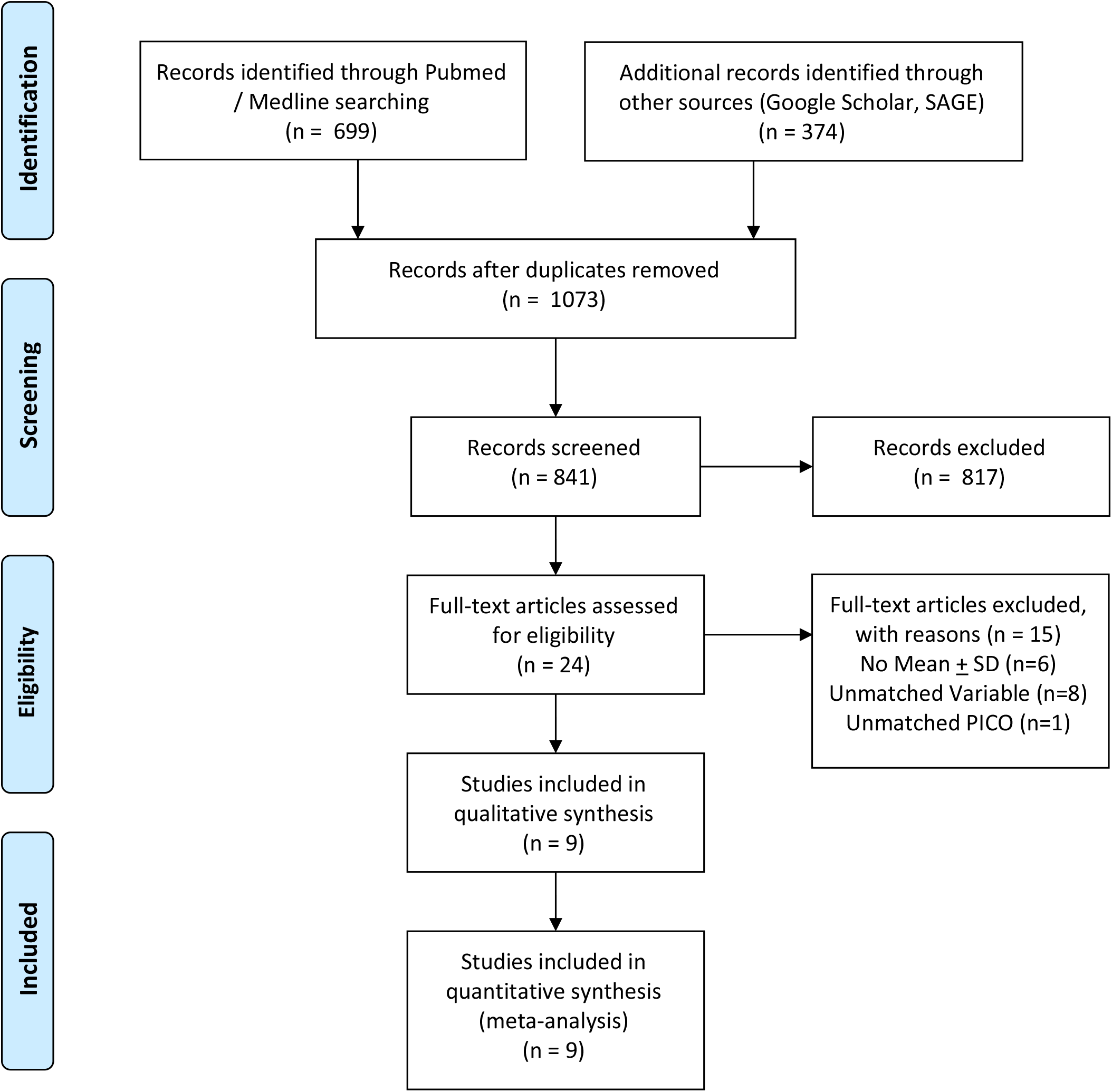
PRISMA flowchart of studies’ selection.

Of the 1073 research articles obtained through search using predefined keywords, nine research articles were obtained that met the inclusion and exclusion criteria and had passed the eligibility test. The nine studies consisted of various ethnicities, races and locations. One article from China, Norway, Canada, the Netherlands, Sweden and Croatia, and three articles from Turkey (Table 1).

**Table 1.** Baseline Characteristics of Studies Included in the Meta-Analysis

| Number | First Author | Year | Journal | Country | N subjects |  | CIMT (mm) |  |
| --- | --- | --- | --- | --- | --- | --- | --- | --- |
|  |  |  |  |  | PE | C | PE | C |
| 1. | Gokhan Goynumer<sup>9</sup> | 2012 | Journal of Clinical Ultrasound | Turkey | 34 | 42 | 0.66 ± 0.07 | 0.62 ± 0.06 |
| 2. | L.-J. Yuan<sup>10</sup> | 2013 | Ultrasound Obstetric Gynecology | China | 22 | 28 | 0.459 ± 0.095 | 0.351 ± 0.085 |
| 3. | Miriam Kristine Sandvik<sup>11</sup> | 2013 | American Journal of Obstetrics and Gynecology | Norway | 89 | 69 | 0.49 ± 0.07 | 0.50 ± 0.06 |
| 4. | Sarah D. Mcdonald<sup>12</sup> | 2013 | Journal of Atherosclerosis | Canada | 100 | 219 | 0.649 ± 0.120 | 0.646 ± 0.111 |
| 5. | Judith Blaauw<sup>13</sup> | 2014 | ACTA Obstetrica et Gynecologica | Netherlands | 17 | 16 | 0.61 ± 0.08 | 0.59 ± 0.06 |
| 6. | Tansim Akhter<sup>8</sup> | 2014 | Ultrasound Obstetric Gynecology | Sweden | 42 | 44 | 0.63 ± 0.12 | 0.61 ± 0.12 |
| 7. | Ciftci Faika Ceylan<sup>14</sup> | 2014 | Journal of the American Society of the Hypertension | Turkey | 46 | 38 | 0.58 ± 0.14 | 0.50 ± 0.10 |
| 8. | Fatma Aykas<sup>15</sup> | 2015 | Journal of Investigative Medicine | Turkey | 25 | 20 | 0.64 ± 0.12 | 0.52 ± 0.08 |
| 9. | Jasna Čerkez Habek<sup>2</sup> | 2018 | Medica Jadertina | Croatia | 55 | 50 | 0.419 ± 0.29 | 0.412 ± 0.43 |

### Funnel Plot and Forest Plot Comparison of CIMT between Women with a History of Preeclampsia and Normal Pregnancy

Four study articles with CIMT differed significantly in the preeclampsia group compared to the control group, while five study articles did not show any significant difference between the two study groups as seen in the Funnel Plot (Figure 2).

**Figure 2.**
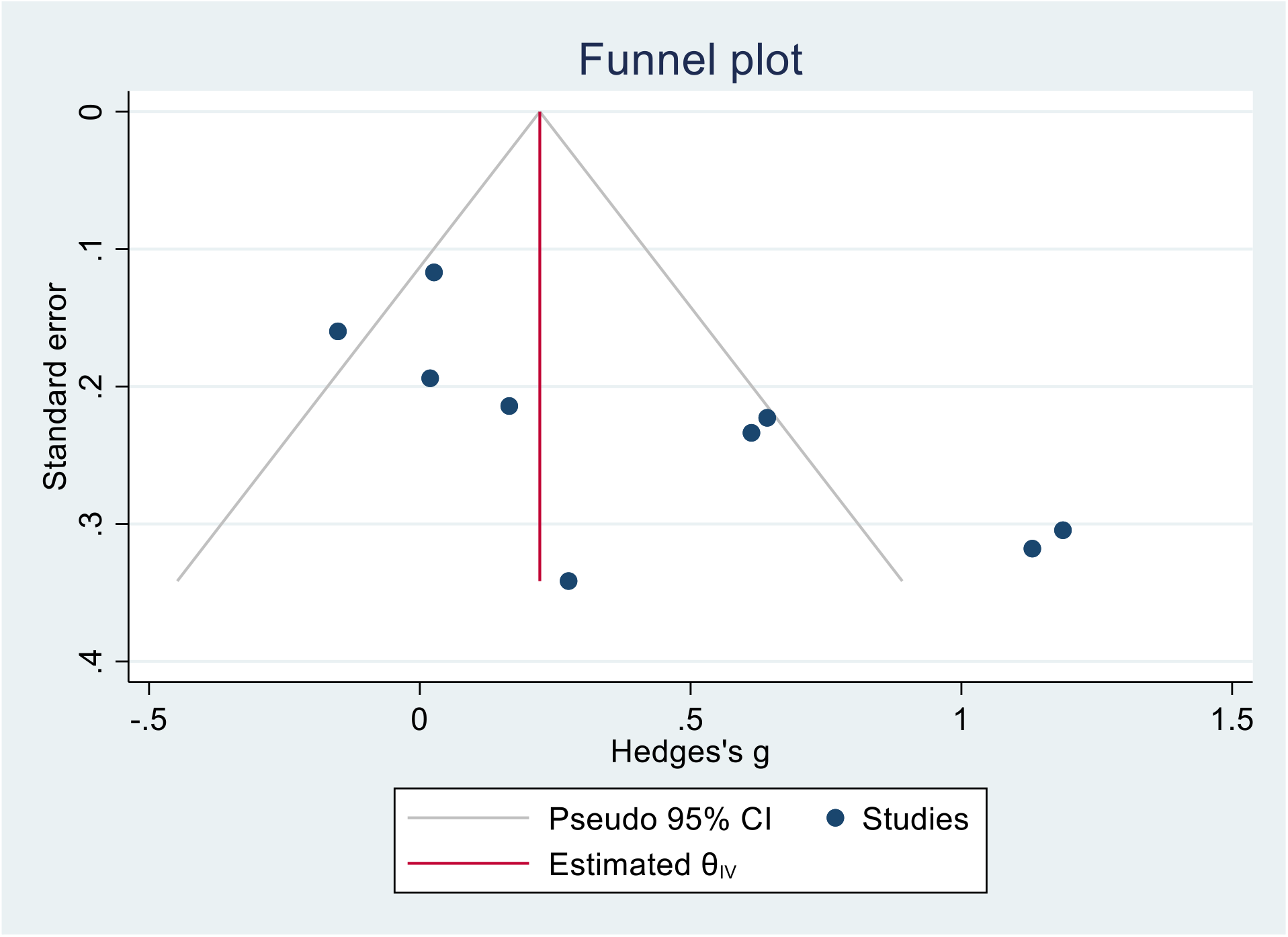
Funnel plot for testing publication bias.

Goynumer et al (2012) performed CIMT examination 1-2 years post-partum in 34 patients with preeclampsia and 42 control patients. CIMT examination was performed on the right and left CCA segments. The mean value of CIMT was 0.66 ± 0.07 mm in the preeclampsia group and 0.62 ± 0.06 in the control group. CIMT in the preeclampsia group was significantly higher than the control group (*p* value 0.025).^9^

Yuan et al (2013) performed a CIMT examination 18 months post-partum in 22 preeclampsia patients and 28 control patients. The CIMT examination was performed on the right CCA segment. The mean value of CIMT was 0.459 ± 0.095 mm in the preeclampsia group and 0.351 ± 0.085 in the control group. CIMT in the preeclampsia group was significantly higher than the control group (*p* value 0.0001).^10^

Sandvik et al (2013) performed a 10-year post-partum CIMT examination in 89 preeclampsia patients and 69 control patients. CIMT examination was performed on the right and left CCA segments. The mean value of CIMT was 0.49 ± 0.07 mm in the preeclampsia group and 0.50 ± 0.06 in the control group. CIMT in the preeclampsia group was not significantly different from the control group (*p* value 0.67).^11^

McDonald et al (2013) performed CIMT examinations 19-28 years post-partum in 109 preeclampsia patients and 219 control patients. CIMT examination was performed on the right and left CCA segments. The mean value of CIMT was 0.649 ± 0.120 mm in the preeclampsia group and 0.646 ± 0.111 in the control group. CIMT on 59 preeclampsia group was not significantly different from the control group (*p* value 1.00).^12^

Blaauw et al (2014) performed CIMT examinations 4-5 years post-partum in 17 preeclampsia patients and 16 control patients. CIMT examination was performed on the right and left CCA segments. The mean value of CIMT was 0.61 ± 0.08 mm in the preeclampsia group and 0.59 ± 0.06 in the control group. CIMT in the preeclampsia group was not significantly different from the control group (*p* value 0.4).^13^

Akhter et al (2014) conducted CIMT examination on 42 preeclampsia patients and 44 control patients aged 40-50 years who were registered in the registry of Uppsala University Hospital, Norway. CIMT examination was performed on the left CCA segment. The mean value of CIMT was 0.63 ± 0.12 mm in the preeclampsia group and 0.61 ± 0.12 in the control group. CIMT in the preeclampsia group was not significantly different from the control group.^8^

Ceylan et al (2014) performed 5 years post-partum CIMT examination in 46 preeclampsia patients and 38 control patients. The CIMT examination was performed on the right CCA segment. The mean value of CIMT was 0.58 ± 0.14 mm in the preeclampsia group and 0.50 ± 0.10 in the control group. CIMT in the preeclampsia group was significantly higher than the control group (*p* value 0.004).^14^

Aykas et al (2015) performed 5 years post-partum CIMT examination in 25 preeclampsia patients and 20 control patients. CIMT examination is performed on the right and left segments. The mean value of CIMT was 0.64 ± 0.12 mm in the preeclampsia group and 0.52 ± 0.08 in the control group. CIMT in the preeclampsia group was significantly higher than the control group (*p* value 0.001).^15^

Habek et al (2018) performed a CIMT examination 6 months post-partum in 55 preeclampsia patients and 50 control patients. CIMT examination was performed on the right and left CCA segments. The mean value of CIMT was 0.419 ± 0.29 mm in the preeclampsia group and 0.412 ± 0.43 in the control group. CIMT in the preeclampsia group was not significantly different compared to the control group.^2^

The combined total mean difference between the studies and their confidence intervals is depicted on the forest plot (Figure 3). The heterogeneity test of the nine research articles was 79.71%, indicating that the nine selected research articles were heterogeneous. The random effect test comparison of CIMT in the preeclampsia and control groups showed p = 0.01 with a confidence interval of 0.08 to 0.70. These results indicate that there is a significant difference between CIMT in the preeclampsia group and the control group, where the preeclampsia group has a greater CIMT value.

**Figure 3.**
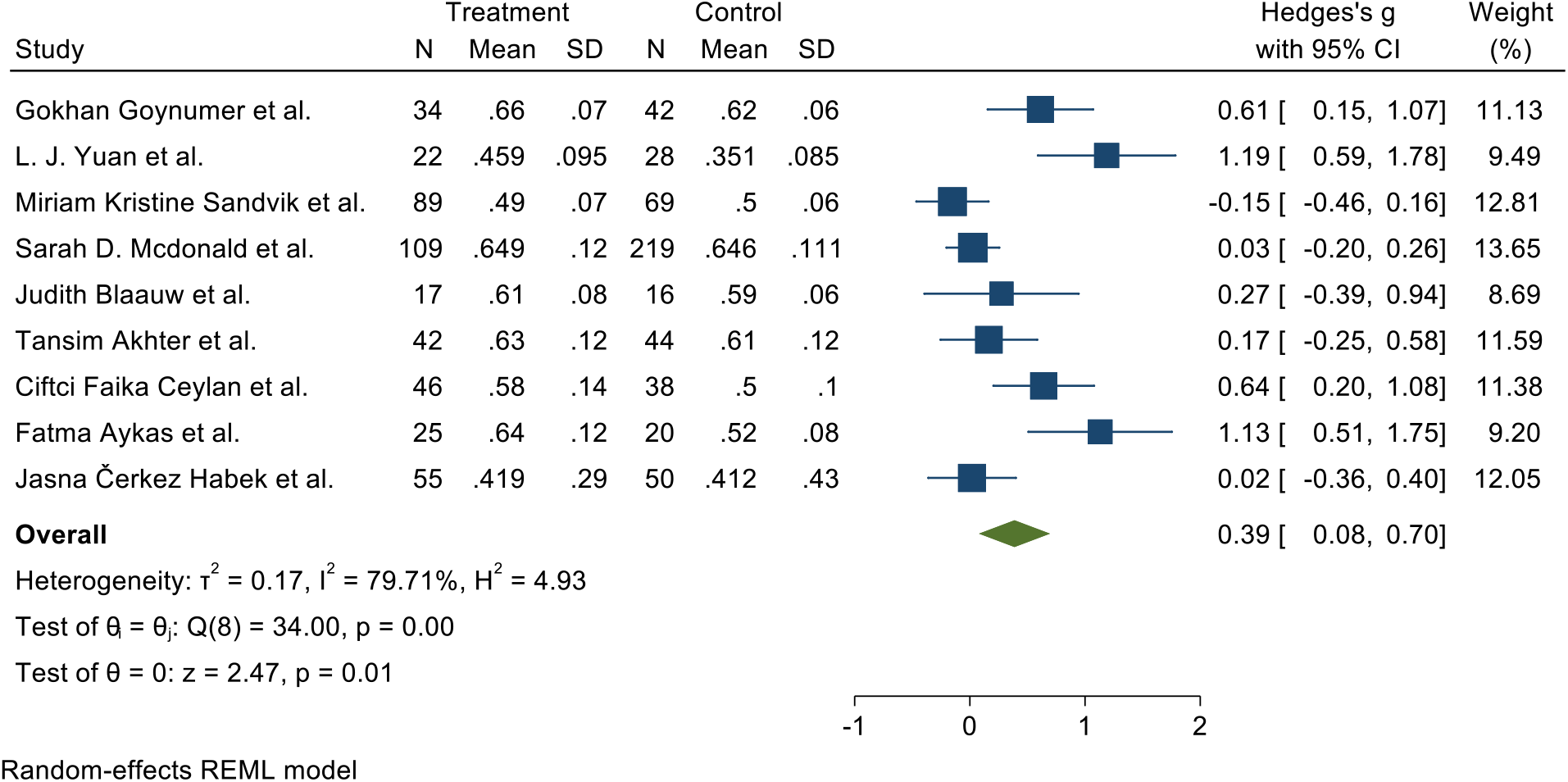
Forest Plot for the effect of CIMT on preeclampsia.

## DISCUSSION

The results of our meta-analysis are supported by the results of two previous meta-analyses regarding subclinical atherosclerosis in preeclamptic patients, namely from Milic et al (2017) and Garovic et al (2017). Milic et al (2017) conducted a meta-analysis of 7 studies that performed CIMT assessments in preeclampsia patients at diagnosis and 10 studies that performed CIMT assessments up to 10 years post-partum. It was concluded that women with preeclampsia had a significantly higher CIMT than women without preeclampsia, both at the time of diagnosis of preeclampsia (SMD: 1.10, 95% CI: 0.73- 1.48, *p* <0.001) and in the first decade post-partum (SMD: 0.58)., 95% CI: 0.36-0.79, *p* <0.001).^16^ Garovic et al (2017) conducted a meta-analysis of 10 studies that performed a CIMT assessment of 10 years or more post-partum. There was a significantly higher CIMT in the preeclampsia group than in the control group (SMD: 0.18, 95% CI: 0.05-0.30, p = 0.004).^17^

Arterial stiffness is a pathological process that develops secondary to changes within the arterial system such as degeneration of elastin and increases in collagen, leading to a thickening of the arterial wall.^18^ CIMT has been widely used to assess atherosclerosis.^19^ However, current guidelines do not support testing CIMT as a risk factor for cardiovascular disease in the general population. This recommendation is based on available evidence that adding a CIMT measurement to the Framingham Risk Score is associated with only a small insignificant increase in the prediction of cardiovascular events at 10 years. On the other hand, recent research on the role of CIMT for risk prediction in certain subclasses has yielded promising results. Meta-analyses of studies in patients with type 1 diabetes, SLE and anti-phospholipid syndrome provide evidence that CIMT may be a good marker of cardiovascular risk in certain populations.^16^ Meanwhile, our meta-analysis indicated that there is a greater atherosclerotic burden as measured by CIMT in patients with a history of preeclampsia. It is this atherosclerosis process that may contribute to cardiovascular complications in patients with a history of preeclampsia.^5,20,21^

### Strength and Limitation of This Review

This study has several limitations, namely the number of research articles that meet the inclusion and exclusion criteria is too small and the sample size of each study is small. The comparison group was only patients with normal pregnancies, excluding patients with chronic hypertension or other spectrum of gestational hypertension. In addition, this meta-analysis study also only included CIMT as a marker of subclinical atherosclerosis, without comparison of other measurement modalities or biomarkers of atherosclerosis.

## CONCLUSION AND PRACTICAL IMPLICATIONS

Carotid intima media thickness in women with a history of preeclampsia is higher than in normal pregnancies. Thus, CIMT examination can be used as a routine examination to stratify the risk of cardiovascular disease in patients with preeclampsia. Further meta-analysis study with a larger scale and broader variables is needed. Any included various modalities and biomarkers to assess the presence of subclinical atherosclerosis in preeclampsia is needed to prove the strong relationship between CIMT and cardiovascular events in women with a history of preeclampsia.

## Data Availability

The datasets generated during and/or analysed during the current study are not publicly available due to protecting participant confidentiality but are available from the corresponding author on reasonable request.

## ACKNOWLEDGEMENT

We would also like to show our gratitude to Johanes Nugroho Eko Putranto for sharing their pearls of wisdom with us during the writing process, and we thank for anonymous residents and staffs for their so-called insights. We are also immensely grateful to Hendri Susilo for his comments on an earlier version of the manuscript, although any errors are our own and should not tarnish the reputations of these esteemed persons.

